# Intensive COVID-19 testing associated with reduced mortality - an ecological analysis of 108 countries

**DOI:** 10.1101/2020.05.28.20115691

**Authors:** Chris Kenyon

## Abstract

**Background:** Intensive screening and testing for COVID-19 could facilitate early detection and isolation of infected persons and thereby control the size of the epidemic. It could also facilitate earlier and more targeted therapy. These factors could plausibly reduce attributable mortality which was the hypothesis tested in this study.

**Methods:** Linear regression was used to assess the country-level association between COVID-19 attributable mortality per 100 000 inhabitants (mortality/capita) and COVID-19 tests/capita (number of tests/100 000 inhabitants) controlling for the cumulative number of COVID-19 infections/100 000 inhabitants (cases/capita), the age of the epidemic (number of days between first case reported and 8 April), national health expenditure per capita and WHO world region.

**Results:** The COVID-19 mortality rate varied between 0.3 and 3110 deaths/100 000 inhabitants (median 30, IQR 8–105). The intensity of testing per 100 000 also varied considerably (median 21,970, IQR 2,735–89,095) as did the number of COVID-19 cases per 100 000 (median 1,600, IQR 340–4,760 cases/100 000). In the multivariate model, the COVID-19 mortality rate was negatively associated with tests/capita (Coef. –0.036, 95% CI –0.047- –0.025) and positively associated with cases/capita (Coef. 0.093, 95% CI 0.819- 1.034).

**Conclusions:** The results are compatible with the hypothesis that intensive testing and isolation could play a role in reducing COVID-10 mortality rates.

## Background

It is important to understand the determinants of national variations in COVID-19-related mortality [1–4]. We aimed to evaluate if testing intensity plays a role. An obvious determinant of COVID-19 attributable mortality is the size of the national epidemic [2, 5, 6]. The measured extent of spread is however heavily dependent on the age of the epidemic and national testing policies [5, 6]. Because a majority of infections are pauci- or a-symptomatic a country that tests less intensively will tend to have a falsely low measured incidence and a correspondingly higher case fatality rate as fewer pauci-symptomatic infections are included in the denominator [5, 7]. Intensive screening and testing could however facilitate early detection and isolation of infected persons and thereby control the size of the epidemic [6, 8]. It could also facilitate earlier and more targeted therapy [8]. These factors could plausibly reduce attributable mortality [6].

We tested this hypothesis by using linear regression to assess the country-level association between COVID-19 attributable mortality per 100 000 inhabitants (mortality/capita) and COVID-19 tests/capita (number of tests/100 000 inhabitants) controlling for the cumulative number of COVID-19 infections/100 000 inhabitants (cases/capita), the age of the epidemic (number of days between first case reported and 8 April), national health expenditure per capita and WHO world region.

## Methods

### Dependent variable

#### Mortality/capita

The COVID-19 attributable mortality per 100 000 inhabitants. This data was obtained from the World of Meters data repository on 8 April 2020: https://www.worldometers.info/coronavirus/

### Independent variables

#### Cases/capita

The cumulative number of cases of COVID-19 infection per 100 000 inhabitants on 8 April 2020 per country. This data was obtained from the World of Meters data repository on 8 April 2020: https://www.worldometers.info/coronavirus/

#### Tests/capita

Cumulative number of nucleic acid amplification SARS CoV-2 tests conducted per country per 100 000 inhabitants up till 8 April 2020. This data was obtained from the World of Meters data repository on 8 April 2020: https://www.worldometers.info/coronavirus/

#### Age of epidemic

The date the first case of COVID-19 was diagnosed in each country. This was measured in days after the first COVID-19 case was officially reported in China (10 January 2020). This data was obtained from the ECDC data repository on 8 April 2020: https://www.ecdc.europa.eu/en/geographical-distribution-2019-ncov-cases

#### Health expenditure per capita

Current expenditures on health per capita in current US dollars for the year 2016. Estimates of current health expenditures include healthcare goods and services consumed during each year. This data was taken from the World Bank’s Data Catalogue: https://datacatalog.worldbank.org/current-health-expenditure-capita-current-us

#### WHO regions

Countries were categorized according to WHO region: https://www.who.int/choice/demography/by_country/en/

### Data analysis

A correlation matrix was performed to investigate the relationships between case fatality rate and the independent variables (Table 1). Linear regression was then used to analyze the association between the COVID-19 mortality rate/100 000 inhabitants and COVID-19 tests/capita (number of tests/100 000 inhabitants) controlling for the cumulative number of COVID-19 infections/100 000 inhabitants (cases/capita), the age of the epidemic (number of days between first case reported and 8 April), national health expenditure per capita and WHO world region. The most recent data available as of 8 April 2020 was used for all variables. Sensitivity analyses were conducted limiting the analysis to countries whose epidemics were older than 15 March 2020. A p-value of < 0.01 was considered statistically significant. The analyses were performed in STATA version 16 (Stata Corp, College Station, Tx).

**Table 1.** Correlation coefficients (r-values) for relations between COVID-19 mortality rate/100 000 inhabitants, cumulative number of cases of COVID-19/100 000, testing rate/100 000, age of the COVID-19 epidemic and health expenditure per capita (* P<0.001)

|  | Mortality | Cumulative Cases | Testing Intensity | Age of Epidemic | Health Expenditure |
| --- | --- | --- | --- | --- | --- |
| Mortality Rate | 1 |  |  |  |  |
| Cumulative Cases | 0.80* | 1 |  |  |  |
| Testing Intensity | 0.17 | 0.55* | 1 |  |  |
| Age of Epidemic | -0.14 | -0.13 | -0.16 | 1 |  |
| Health Expenditure | 0.27 | 0.51* | 0.45* | -0.43* | 1 |

## Results

Complete data was available for 108 countries. The COVID-19 mortality rate varied between 0.3 and 3110 deaths/100 000 inhabitants (median 30, IQR 8–105; Table 3). The intensity of testing per 100 000 also varied considerably (median 21,970, IQR 2,735–89,095) as did the number of COVID-19 cases per 100 000 (median 1,600, IQR 340–4,760 cases/100 000)

Pearson’s correlation revealed deaths/capita to be positively correlated with cases/capita (r = 0.80, P<0.001), which was in turn positively correlated with tests/capita (r = 0.55, P<0.001), and health care expenditure (r = 0.51, P<0.001; Table 1). Tests/capita was positively correlated with health care expenditure (r = 0.45, P<0.001).

In the multivariate model, the COVID-19 mortality rate was negatively associated with tests/capita (Coef. –0.036, 95% CI –0.047- –0.025) and positively associated with cases/capita (Coef. 0.093, 95% CI 0.819- 1.034; Table 2). Sensitivity analyses limited to countries with epidemics older than 15 March 2020 made little difference to the findings (Table 4).

**Table 2.**
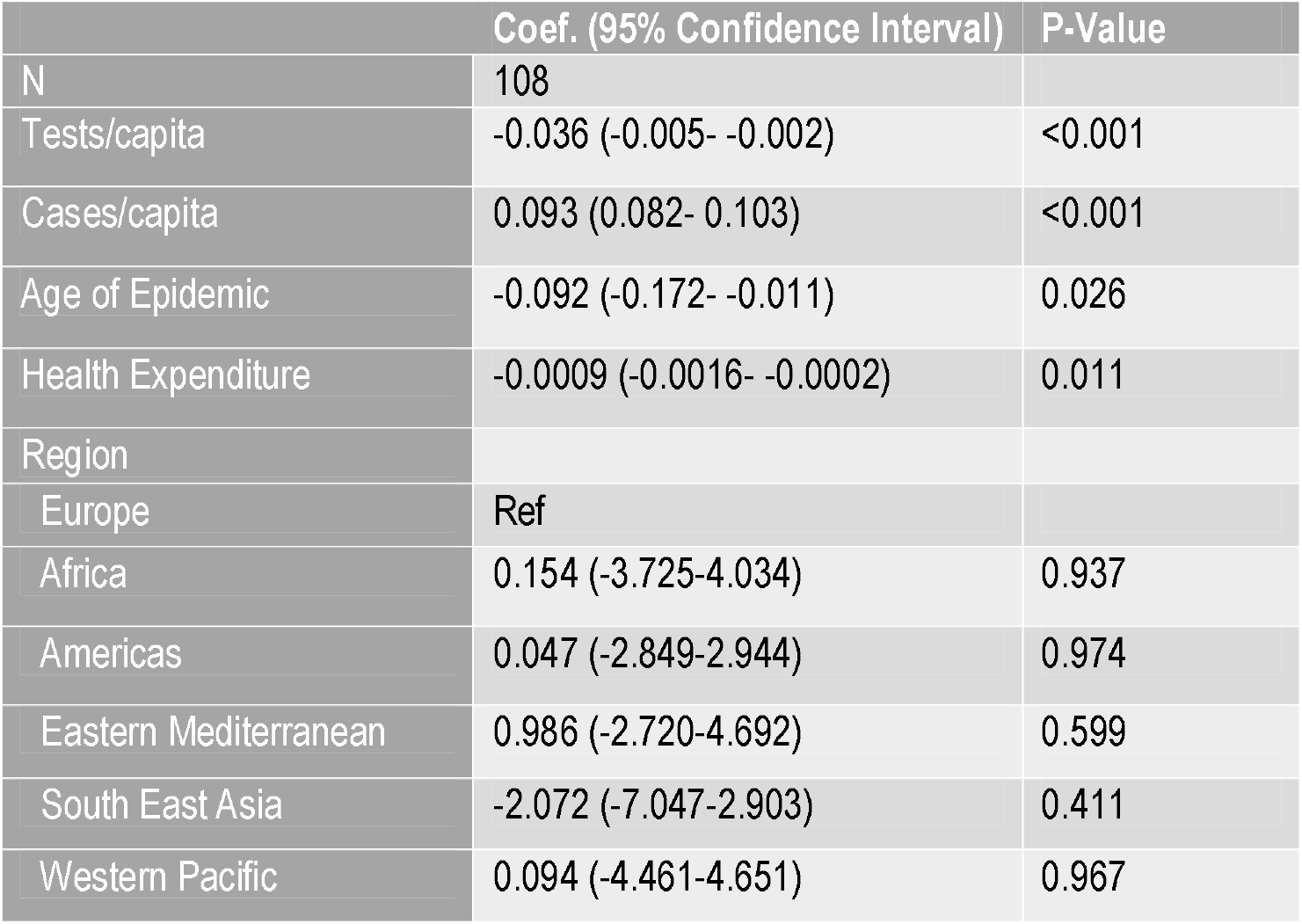
Country level, multivariate linear regression of factors associated with COVID-19 mortality rate per 100 000 inhabitants.

**Table 3.** COVID-19 mortality rate/100 000 inhabitants, cumulative number of cases of COVID-19/100 000 (Cases/capita), number tested for COVID-19/100 000 (Tests/capita), date first COVID-19 case reported (Age Epi.) and health expenditure (Health Exp – in US$) in 108 countries.

| Country | Mortality | Cases/capita | Tests/Capita | Age Epi. | Health Exp. |
| --- | --- | --- | --- | --- | --- |
| Albania | 0.8 | 13.9 | 103.9 | 07-Mar-20 | 272 |
| Algeria | 0.4 | 3.3 | 7.7 | 24-Feb-20 | 260 |
| Andorra | 29.8 | 730 | 2165.3 | 01-Mar-20 | 3835 |
| Antigua and Barbuda | 2 | 19.4 | 40.8 | 12-Mar-20 | 623 |
| Argentina | 0.1 | 3.8 | 26.1 | 02-Mar-20 | 955 |
| Armenia | 0.3 | 29.7 | 184 | 29-Feb-20 | 359 |
| Australia | 0.2 | 23.6 | 1252.4 | 24-Jan-20 | 5002 |
| Austria | 3 | 143.2 | 1340.8 | 24-Feb-20 | 4688 |
| Azerbaijan | 0.08 | 8.1 | 525.7 | 27-Feb-20 | 268 |
| Bahrain | 0.3 | 48.2 | 3076.6 | 23-Feb-20 | 1099 |
| Bangladesh | 0.01 | 0.1 | 2.6 | 07-Mar-20 | 34 |
| Barbados | 1 | 21.9 | 193.8 | 16-Mar-20 | 1164 |
| Belarus | 0.1 | 11.3 | 486.8 | 27-Feb-20 | 318 |
| Belgium | 19.3 | 201.9 | 726.9 | 03-Feb-20 | 4149 |
| Belize | 0.3 | 2 | 67.4 | 22-Mar-20 | 304 |
| Bolivia | 0.1 | 1.8 | 5.1 | 09-Mar-20 | 213 |
| Bosnia and Herzegovina | 1 | 24.5 | 210.6 | 04-Mar-20 | 444 |
| Brazil | 0.3 | 6.7 | 25.8 | 24-Feb-20 | 1016 |
| Brunei | 0.2 | 30.9 | 2021.8 | 08-Mar-20 | 630 |
| Bulgaria | 0.3 | 8.5 | 228.8 | 06-Mar-20 | 612 |
| Canada | 1 | 48.9 | 922.3 | 24-Jan-20 | 4458 |
| Chile | 0.3 | 29 | 315.9 | 02-Mar-20 | 1191 |
| Colombia | 0.1 | 3.5 | 59.8 | 05-Mar-20 | 340 |
| Costa Rica | 0.04 | 9.5 | 108.5 | 04-Mar-20 | 889 |
| Croatia | 0.5 | 32.7 | 319.7 | 24-Feb-20 | 884 |
| Cuba | 0.1 | 3.5 | 62.3 | 10-Mar-20 | 971 |
| Cyprus | 0.7 | 43.6 | 1182.2 | 08-Mar-20 | 1634 |
| Czechia | 0.8 | 47 | 921.5 | 29-Feb-20 | 1321 |
| Denmark | 3.8 | 93 | 1008.6 | 26-Feb-20 | 5566 |
| Dominican Republic | 1 | 19.5 | 41.9 | 29-Feb-20 | 414 |
| Ecuador | 1.2 | 22.6 | 81.7 | 28-Feb-20 | 505 |
| Egypt | 0.09 | 1.4 | 24.4 | 13-Feb-20 | 131 |
| Estonia | 1.8 | 89.3 | 1870.5 | 26-Feb-20 | 1185 |
| Ethiopia | 0.002 | 0.05 | 2.2 | 12-Mar-20 | 28 |
| Finland | 0.7 | 44.9 | 658.8 | 28-Jan-20 | 4117 |
| France | 15.8 | 167.1 | 343.6 | 23-Jan-20 | 4263 |
| Georgia | 0.08 | 5.2 | 69.2 | 25-Feb-20 | 308 |
| Germany | 2.5 | 130.5 | 1096.2 | 26-Jan-20 | 4714 |
| Greece | 0.8 | 18.1 | 274.2 | 25-Feb-20 | 1511 |
| Guatemala | 0.02 | 0.5 | 6.3 | 12-Mar-20 | 241 |
| Guyana | 0.6 | 4.2 | 16.8 | 11-Mar-20 | 192 |
| Haiti | 0.009 | 0.2 | 2.3 | 19-Mar-20 | 38 |
| Hungary | 0.6 | 9.3 | 266.5 | 03-Mar-20 | 943 |
| Iceland | 1.8 | 473.6 | 9068.9 | 27-Feb-20 | 5064 |
| India | 0.01 | 0.4 | 10.2 | 29-Jan-20 | 63 |
| Indonesia | 0.09 | 1.1 | 5.2 | 01-Mar-20 | 112 |
| Iran | 4.8 | 76.9 | 251.4 | 18-Feb-20 | 415 |
| Iraq | 0.2 | 3 | 70.6 | 21-Feb-20 | 153 |
| Ireland | 4.3 | 115.6 | 860.4 | 28-Feb-20 | 4759 |
| Israel | 0.8 | 108.6 | 1355.7 | 20-Feb-20 | 2837 |
| Italy | 28.3 | 224.3 | 1249.5 | 29-Jan-20 | 2739 |
| Jamaica | 0.1 | 2.1 | 18.2 | 09-Mar-20 | 296 |
| Japan | 0.07 | 3.4 | 43.7 | 14-Jan-20 | 4233 |
| Jordan | 0.06 | 3.5 | 166.6 | 01-Mar-20 | 224 |
| Kazakhstan | 0.04 | 3.8 | 302.4 | 12-Mar-20 | 262 |
| Kenya | 0.01 | 0.3 | 9.2 | 12-Mar-20 | 66 |
| Kyrgyzstan | 0.06 | 4.1 | 147.4 | 17-Mar-20 | 73 |
| Latvia | 0.1 | 30.6 | 1274.3 | 01-Mar-20 | 874 |
| Lebanon | 0.3 | 8.4 | 158.3 | 20-Feb-20 | 662 |
| Lithuania | 0.6 | 33.5 | 1106 | 27-Feb-20 | 988 |
| Luxembourg | 7 | 474.5 | 4105.9 | 28-Feb-20 | 6271 |
| Malaysia | 0.2 | 12.7 | 179.9 | 24-Jan-20 | 362 |
| Mauritania | 0.02 | 0.1 | 1.4 | 12-Mar-20 | 47 |
| Mauritius | 0.6 | 21.5 | 386.3 | 17-Mar-20 | 553 |
| Mexico | 0.1 | 2.2 | 19.7 | 27-Feb-20 | 462 |
| Moldova | 0.7 | 29.1 | 126.6 | 06-Mar-20 | 171 |
| Montenegro | 0.3 | 39.5 | 320.7 | 16-Mar-20 | 532 |
| Morocco | 0.2 | 3.4 | 15.2 | 01-Mar-20 | 171 |
| Myanmar | 0.006 | 0.04 | 2.3 | 22-Mar-20 | 62 |
| Netherlands | 13.1 | 119.9 | 505.3 | 26-Feb-20 | 4742 |
| New Zealand | 0.02 | 25.1 | 972.1 | 27-Feb-20 | 3745 |
| Niger | 0.05 | 1.1 | 17.3 | 18-Mar-20 | 23 |
| Nigeria | 0.003 | 0.1 | 2.4 | 27-Feb-20 | 79 |
| North Macedonia | 1.4 | 29.6 | 299 | 25-Feb-20 | 328 |
| Norway | 1.7 | 112.3 | 2100.9 | 25-Feb-20 | 7478 |
| Pakistan | 0.03 | 1.9 | 19.1 | 25-Feb-20 | 40 |
| Panama | 1.4 | 52.1 | 238.6 | 09-Mar-20 | 1041 |
| Paraguay | 0.07 | 1.7 | 28.6 | 06-Mar-20 | 327 |
| Peru | 0.3 | 9 | 65.4 | 05-Mar-20 | 316 |
| Philippines | 0.2 | 3.5 | 22.4 | 29-Jan-20 | 129 |
| Poland | 0.4 | 13.2 | 262.3 | 03-Mar-20 | 809 |
| **Portugal** | 3.7 | 128.9 | 1263 | 01-Mar-20 | 1801 |
| **Qatar** | 0.2 | 76.7 | 1451.5 | 28-Feb-20 | 1827 |
| **Romania** | 1.1 | 24.7 | 245.4 | 25-Feb-20 | 476 |
| **Russia** | 0.04 | 5.9 | 623.7 | 30-Jan-20 | 469 |
| **San Marino** | 100.2 | 822.3 | 1727 | 26-Feb-20 | 3013 |
| **Serbia** | 0.7 | 30.5 | 123.2 | 05-Mar-20 | 494 |
| **Singapore** | 0.1 | 27.7 | 1111 | 22-Jan-20 | 2462 |
| **Slovakia** | 0.04 | 12.5 | 360.5 | 05-Mar-20 | 1178 |
| **Slovenia** | 1.9 | 52.5 | 1475.2 | 03-Mar-20 | 1834 |
| **South Africa** | 0.02 | 2.9 | 98 | 04-Mar-20 | 428 |
| **South Korea** | 0.4 | 20.3 | 931 | 19-Jan-20 | 2043 |
| **Spain** | 31.1 | 313.7 | 759.3 | 30-Jan-20 | 2390 |
| **Sri Lanka** | 0.03 | 0.9 | 15.2 | 26-Jan-20 | 153 |
| **Sweden** | 6.8 | 83.4 | 541.6 | 30-Jan-20 | 5711 |
| **Switzerland** | 9.9 | 263.3 | 1986.7 | 24-Feb-20 | 9836 |
| **Thailand** | 0.04 | 3.4 | 103 | 12-Jan-20 | 222 |
| **Togo** | 0.04 | 0.8 | 21.1 | 05-Mar-20 | 39 |
| **Trinidad and Tobago** | 0.6 | 7.6 | 62.7 | 11-Mar-20 | 1064 |
| **Tunisia** | 0.2 | 5.3 | 70 | 01-Mar-20 | 257 |
| **Turkey** | 0.9 | 40.4 | 264.3 | 09-Mar-20 | 469 |
| **Ukraine** | 0.1 | 3.8 | 16.5 | 02-Mar-20 | 141 |
| **United Arab Emirates** | 0.1 | 26.9 | 5451.7 | 28-Jan-20 | 1323 |
| **United Kingdom** | 9.1 | 81.4 | 392.9 | 30-Jan-20 | 3958 |
| **United States** | 3.9 | 122.1 | 636.7 | 20-Jan-20 | 9870 |
| **Uruguay** | 0.2 | 12.2 | 164.9 | 12-Mar-20 | 1379 |
| **Zambia** | 0.005 | 0.2 | 3.4 | 17-Mar-20 | 57 |
| **Zimbabwe** | 0.01 | 0.07 | 2.5 | 19-Mar-20 | 94 |

**Table 4.** Country level, multivariate linear regression of factors associated with COVID-19 mortality rate per 100 000 inhabitants. Sensitivity analysis limited to countries with epidemics starting prior to 15 March 2020.

|  | Coef. (95% Confidence Interval) | P-Value |
| --- | --- | --- |
| N | 98 |  |
| Tests/capita | -0.036 (-0.005- -0.002) | <0.001 |
| Cases/capita | 0.093 (0.081- 0.104) | <0.001 |
| Age of Epidemic | -0.107 (-0.200- -0.014) | 0.024 |
| Health Expenditure | -0.0010 (-0.0017- -0.0002) | 0.012 |
| Region |  |  |
| Europe | Ref |  |
| Africa | -0.163 (-4.992-4.664) | 0.946 |
| Americas | 0.046 (-3.249-3.149) | 0.977 |
| Eastern Mediterranean | 0.906 (-3.0188-4.830) | 0.648 |
| South East Asia | -3.037(-8.874-2.799) | 0.304 |
| Western Pacific | 0.240 (-5.091-4.651) | 0.922 |

## Conclusion

Our analysis confirms the logical association between increased testing intensity and increased detection of cases/capita as well the association between cases/capita and deaths/capita. The key finding was that on multivariate analysis, testing intensity was negatively associated with mortality/capita. This is compatible with the theory that intensive testing is associated with reduced mortality via reducing the spread of SARS-CoV-2 and/or facilitating earlier more appropriate treatment of infected persons [6, 8]. Our study design does not allow us to exclude the possibility that this association is due to residual confounding. Possible confounders include other components of a successful national COVID-19 containment strategy such as contact tracing or social distancing [2, 5]. These may be associated with both increased testing intensity and reduced transmission/deaths. In addition, we did not control for other variables such as national age structure and prevalence of comorbidities which may have influenced mortality [9].

These limitations notwithstanding, our analysis provides additional evidence to promote calls to intensify national COVID-19 testing to not only control the spread of this disease but also to reduce associated mortality.

## Data Availability

The data used is freely available from the World of Meters data repository: https://www.worldometers.info/coronavirus/ and ECDC: https://www.ecdc.europa.eu/en/geographical-distribution-2019-ncov-cases

https://www.worldometers.info/coronavirus/

## Authors’ contributions

CK conceptualized the study, was responsible for the acquisition, analysis and interpretation of data and wrote the analysis up as a manuscript.

## Ethical approval

The analysis involved a secondary analysis of public access ecological level data. As a result, no ethics approval was necessary.

## Informed consent

Not applicable

## Acknowledgements

Nil

